# Healthcare disparities among anticoagulation therapies for severe COVID-19 patients in the multi-site VIRUS registry

**DOI:** 10.1101/2020.11.06.20226035

**Authors:** Christian Kirkup, Colin Pawlowski, Arjun Puranik, Ian Conrad, John C. O’Horo, Dina Gomaa, Valerie M. Banner-Goodspeed, Jarrod M Mosier, Igor Borisovich Zabolotskikh, Steven K. Daugherty, Michael A. Bernstein, Howard A. Zaren, Vikas Bansal, Brian Pickering, Andrew D. Badley, Rahul Kashyap, AJ Venkatakrishnan, Venky Soundararajan

## Abstract

COVID-19 patients are at an increased risk of thrombosis and various anticoagulants are being used in patient management without an established standard-of-care. Here, we analyze hospitalized and ICU patient outcomes from the *Viral Infection and Respiratory illness Universal Study (VIRUS)* registry. We find that severe COVID patients administered unfractionated heparin but not enoxaparin have a higher mortality-rate (311 deceased patients out of 760 total patients = 41%) compared to patients administered enoxaparin but not unfractionated heparin (214 deceased patients out of 1,432 total patients = 15%), presenting a risk ratio of 2.74 (95% C.I.: [2.35, 3.18]; p-value: 1.4e-41). This difference persists even after balancing on a number of covariates including: demographics, comorbidities, admission diagnoses, and method of oxygenation, with an amplified mortality rate of 39% (215 of 555) for unfractionated heparin vs. 23% (119 of 522) for enoxaparin, presenting a risk ratio of 1.70 (95% C.I.: [1.40, 2.05]; p-value: 2.5e-7). In these balanced cohorts, a number of complications occurred at an elevated rate for patients administered unfractionated heparin compared to those administered enoxaparin, including acute kidney injury (227 of 642 [35%] vs. 156 of 608 [26%] respectively, adjusted p-value 0.0019), acute cardiac injury (40 of 642 [6.2%] vs. 15 of 608 [2.5%] respectively, adjusted p-value 0.01), septic shock (118 of 642 [18%] vs. 73 of 608 [12%] respectively, adjusted p-value 0.01), and anemia (81 of 642 [13%] vs. 46 of 608 [7.6%] respectively, adjusted p-value 0.02). Furthermore, a higher percentage of Black/African American COVID patients (375 of 1,203 [31%]) were noted to receive unfractionated heparin compared to White/Caucasian COVID patients (595 of 2,488 [24%]), for a risk ratio of 1.3 (95% C.I.: [1.17, 1.45], adjusted p-value: 1.6e-5). After balancing upon available clinical covariates, this difference in anticoagulant use remained statistically significant (272 of 959 [28%] for Black/African American vs. 213 of 959 [22%] for White/Caucasian, adjusted p-value: 0.01, relative risk: 1.28, 95% C.I.: [1.09, 1.49]). While retrospective studies cannot suggest any causality, these findings motivate the need for follow-up prospective research in order to elucidate potential socioeconomic, racial, or other disparities underlying the use of anticoagulants to treat severe COVID patients.

## Introduction

Major complications of severe COVID-19 include coagulopathy and cardiovascular events^1–3^. Through the National Institutes of Health (NIH) ACTIV initiative, there are multiple ongoing research studies to evaluate the safety and effectiveness of various types and doses of anticoagulants^4^. According to NIH Director Francis S. Collins, M.D., Ph.D., “There is currently no standard of care for anticoagulation in hospitalized COVID-19 patients, and there is a desperate need for clinical evidence to guide practice.”^4^ Due to the current knowledge gap in evidence-based anticoagulant treatments for severe COVID-19, there are many open questions on topics including: types of anticoagulant medications to prescribe, dosing for anticoagulants, indications for anticoagulant prescriptions, and prophylactic vs. therapeutic use.

In this paper, we focus on which types of anticoagulant medications to prescribe for patients with severe COVID-19. We conduct this analysis on the Society for Critical Care Medicine’s VIRUS registry^5^, a large-scale, international, multi-site study of hospitalized COVID-19 patients. We consider three categories of anticoagulant medications: (1) Unfractionated Heparin, (2) Enoxaparin, and (3) Other types of Low Molecular Weight Heparin (LMWH). First, we consider head-to-head comparisons of enoxaparin vs. unfractionated heparin and enoxaparin vs. other types of LMWH by constructing cohorts of hospitalized COVID patients who received one anticoagulant medication but not the other during their hospital stay for COVID-19. For each cohort comparison, we evaluate patient outcomes including: mortality at hospital discharge, 28-day mortality status, average hospital length of stay in days, average ICU length of stay in days, and complications during the 28-day follow-up period. In addition, for each comparison we repeat the analysis using propensity score matching to control for potential confounding variables including: demographics, comorbidities, evidence of infiltrates, ICU admission status, initial oxygenation method, admission diagnoses, and time in days to anticoagulant administration. Finally, we analyzed the rates of anticoagulant medication administration by race, focusing on cohorts of Black/African American and White/Caucasian patients. Similar, we used propensity score matching to construct race-based cohorts balanced on the clinical covariates listed previously, and we report patient outcomes for both the original and the propensity-matched race-stratified cohorts.

## Methods

### Study Design

The Society of Critical Care Medicine (SCCM)’s *Discovery Viral Infection and Respiratory Illness Universal Study (VIRUS): COVID-19 Registry* is composed of data collected from patients hospitalized for COVID-19. The registry was granted exempt status for human subjects research by the institutional review board at Mayo Clinic (IRB:20-002610). The ClinicalTrials.gov number is NCT04323787. Each study site submitted a proposal to their local review boards for approval and signed a data use agreement before being granted permission to extract and enter de-identified data into the registry.

As of September 26, 2020, the total size of the study population is 28,964 patients reported by 244 hospitals across 21 countries. While a portion of sites report data for each day in the hospital for each patient, emphasis is placed on capturing data at key events in the treatment process. These include the day of admission to the hospital, the first three days in the hospital, and first day of admission to the ICU (if admitted) as well as outcomes measures like the duration of stay in the hospital and the ICU (if admitted) and the 28-day survival status. Data completeness of the features is variable depending on the frequency of updates from the sites. Data for the registry is collected via REDCap and can be automatically filled from a site’s EHR data.

Features reported include comorbidities listed in the VIRUS questionnaire (obesity, diabetes, hypertension, etc.), complications (acute kidney injury, deep vein thrombosis, coagulopathy, etc.), medications prescribed in hospital (antibacterials, anticoagulants, statins, etc.) as well as more refined medication features within a category (*Antivirals*: Remdesivir, Ritonavir, Lopinavir, etc.). Other features collected for each patient include Hospital Length of Stay, ICU Length of Stay, Height, Weight, etc.

Prior studies suggest that enoxaparin may be more efficacious than unfractionated heparin in the treatment of conditions like acute coronary syndromes^6^ and these are two most frequently administered anticoagulants (**Supplementary Table S1**). Thus, we compare the outcomes of patients taking enoxaparin and heparin by constructing two cohorts: (i) patients who were administered enoxaparin but not unfractionated heparin and (ii) patients who were administered unfractionated heparin but not enoxaparin. The cohort sizes were 1814 and 887 respectively. Statistical tests were applied to 21 outcomes (with Benjamini-Hochberg procedure applied to account for the problem of multiple comparisons; details below). Mortality at hospital discharge was the primary outcome of interest. Outcomes that were compared include (1) mortality at hospital discharge, (2) mortality at 28 days, (3) admission to ICU (within 28 days of hospitalization), (4) length of stay in ICU (among alive patients), (5) length of stay in hospital (among alive patients), and the following 16 complications: (6) acute cardiac injury, (7) acute kidney injury, (8) anemia, (9) bacteremia, (10) bacterial pneumonia, (11) cardiac arrest, (12) cardiac arrhythmia, (13) co- or secondary infection, (14) congestive heart failure, (15) deep vein thrombosis, (16) hyperglycemia, (17) liver dysfunction, (18) pleural effusions, (19) ARDS, (20) septic shock, (21) stroke/cerebrovascular incident. The diagnostic criteria available are outlined in **Supplementary Table S2**.

To account for potentially confounding variables, we performed propensity score matching to balance covariates between the two cohorts. The statistical tests for differences in outcomes were repeated on the matched cohorts. The covariates which were balanced include demographics, comorbidities and various features on admission. Further detail on the procedure, including a listing of covariates used, is below. The code to process the raw data files was written in R v3.6.1. The code to perform the statistical analyses was written in Python v3.7.7, using the scikit-learn package v0.23.2 to train the logistic regression models for the propensity score matching step. In **Supplementary Table S3**, we show the data completeness for the clinical covariates that we used for matching. Most covariates have close to full completeness (over 90%), with the exception of the “evidence of infiltrates” covariate, which has roughly 80% data completeness. For this field, missing values were imputed to be the mean of other values of the field within the treatment group.

### Statistical Methods

For each of the cohort comparisons, we ran a series of statistical significance tests in order to compare across each of the patient outcome variables of interest. For categorical outcome variables (e.g. mortality status, complications), we report the proportion of patients in each cohort that have the outcome variable, the relative risk (ratio of proportions for each cohort), 95% confidence interval for the relative risk, and Chi-squared p-value. The function `stats.chi2_contingency` from the SciPy package in Python was used to compute the Chi-squared p-values. For continuous outcome variables (e.g. hospital/ICU length of stay), we report the mean and standard deviation of the variable in each cohort, along with the p-value from a two-sided Mann-Whitney test (`stats.mannwhitneyu` from SciPy) comparing the two cohorts. Finally, we apply the Benjamini-Hochberg correction to adjust p-values for multiple comparisons.

### Propensity score matching

In order to control for potential confounding factors which may explain differences in patient outcomes between the Enoxaparin and Unfractionated Heparin cohorts, we used Propensity Score Matching to balance the cohorts^7^. First, propensity scores for each of the patients in the two cohorts were computed by fitting a logistic regression model as a function of the clinical covariates (listed below). Next, patients from the Enoxaparin and Unfractionated Heparin cohorts were matched using a 1:1 matching ratio and a heuristic caliper of 0.1 x pooled standard deviation^8^, allowing for drops. Prior to matching, there were 1,814 patients in the enoxaparin cohort (administered enoxaparin but not unfractionated heparin), and there were 887 patients in the unfractionated heparin cohort (administered unfractionated heparin but not enoxaparin). From these two cohorts, 659 matched pairs were found, and statistical analyses were run on the final matched cohorts. Here is the full list of covariates that were considered for the propensity score matching step:

- **Demographics:** Age, gender, race, ethnicity.
- **Comorbidities:** Pre-existing conditions, including: (1) asthma, (2) blood loss anemia, (3) cardiac arrhythmias, (4) chronic kidney disease, (5) chronic dialysis, (6) chronic pulmonary disease, (7) coagulopathy, (8) congestive heart failure, (9) coronary artery disease, (10) dementia, (11) depression, (12) diabetes, (13) dyslipidemia/hyperlipidemia, (14) HIV / AIDS or other immunosuppression, (15) hematologic malignancy, (16) Hepatitis B, (17) Hepatitis C, (18) history of solid organ or bone marrow transplant, (19) hypertension, (20) hypothyroidism, (21) iron deficiency anemia, (22) liver disease, (23) malnutrition, (24) metastatic cancer, (25) obstructive sleep apnea / home CPAP / BiPAP use, (26) obesity, (27) paralysis, (28) peptic ulcer disease excluding bleeding, (29) psychosis, (30) pulmonary circulation disorder, (31) rheumatoid arthritis/collagen vascular disease, (32) solid tumor without metastasis, (33) stroke or other neurological disorders, (34) substance use disorder, (35) valvular heart disease, (36) venous thromboembolism (deep vein thrombosis / pulmonary embolism)
- **In ICU on admission to hospital**
- **Admission diagnoses:** Conditions which are present upon admission to hospital for COVID-19, including: (1) acute respiratory distress syndrome (ARDS), (2) non-ARDS acute hypoxic respiratory failure, (3) acute liver injury, (4) acute myocardial infarction, (5) acute renal failure/injury (with or without hemofiltration) (6) bacteremia, (7) bacterial pneumonia, (8) cardiac arrest, (9) cardiac arrhythmias (atrial fibrillation, heart block, torsades des point, ventricular tachycardia), (10) congestive heart failure / cardiomyopathy, (11) delirium / encephalopathy, (12) disseminated intravascular coagulation, (13) gastrointestinal hemorrhage, (14) hyperglycemia, (15) hypoglycemia, (16) meningitis / encephalitis, (17) myocarditis, (18) pleural effusion, (19) pneumothorax, (20) rhabdomyolysis / myositis, (21) seizure, (22) sepsis, (23) shock, (24) stroke.
- **Evidence of infiltrates via X-ray or CT scan**
- **Oxygenation-related features on admission:** Supplemental oxygenation method on day of admission, among: (1) invasive mechanical ventilation, (2) noninvasive ventilation (CPAP or BIPAP), (3) high flow nasal cannula, (4) bag mask oxygen therapy, (5) non-rebreather mask oxygen therapy, (6) nasal cannula, (7) miscellaneous other form of oxygenation.
- **Day of anticoagulant administration:** First day that the patient received the anticoagulant of interest (unfractionated heparin, enoxaparin, or other LMWH), relative to the day of hospital admission for COVID-19.

The same propensity score matching procedure was done with enoxaparin vs. other low molecular weight heparin in place of enoxaparin vs. unfractionated heparin. Propensity score matching was also applied to balance covariates between the Black/African American and White/Caucasian patient cohorts; the “outcome” compared in this case was the rate of administration of each anticoagulant. All of the same covariates (except race and day of anticoagulant administration) listed above were used in this balancing.

## Results

In **Figure 1**, we present the mortality rate and ICU admission rate for patients in the SCCM VIRUS registry^5^ with outcomes data available. Among the 28,964 patients in the VIRUS registry at the time of the study, hospital discharge status was available for 8,623 patients, of which 6,208 (72%) were alive at discharge. For patients that were administered enoxaparin but not unfractionated heparin, hospital discharge status was available for 1,432 patients, of which 1,217 (85%) were alive at discharge. For patients that were administered unfractionated heparin but not enoxaparin, hospital discharge status was available for 760 patients, and 448 (59%) were alive at discharge. Comparing the mortality outcomes (unadjusted), patients in the heparin cohort have a higher mortality rate compared to those in the enoxaparin cohort (risk ratio: 2.74; 95% C.I.: [2.35, 3.18]; p-value: 1.4e-41) (**Figure 1a**). For patients that were administered enoxaparin but not unfractionated heparin, ICU admission status was available for 1,690 patients, of which 794 (47%) were admitted to the ICU. Similarly, for patients that were administered unfractionated heparin but not enoxaparin, ICU admission status was available for 863 patients, of which 570 (66%) were admitted to the ICU. Comparing the ICU admission status (unadjusted), patients administered unfractionated heparin had a higher rate of admission to the ICU compared to patients administered enoxaparin (risk ratio: 1.41; 95% C.I.: [1.32, 1.51]; p-value: 3.4e-20) (**Figure 1b**).

**Figure 1.**
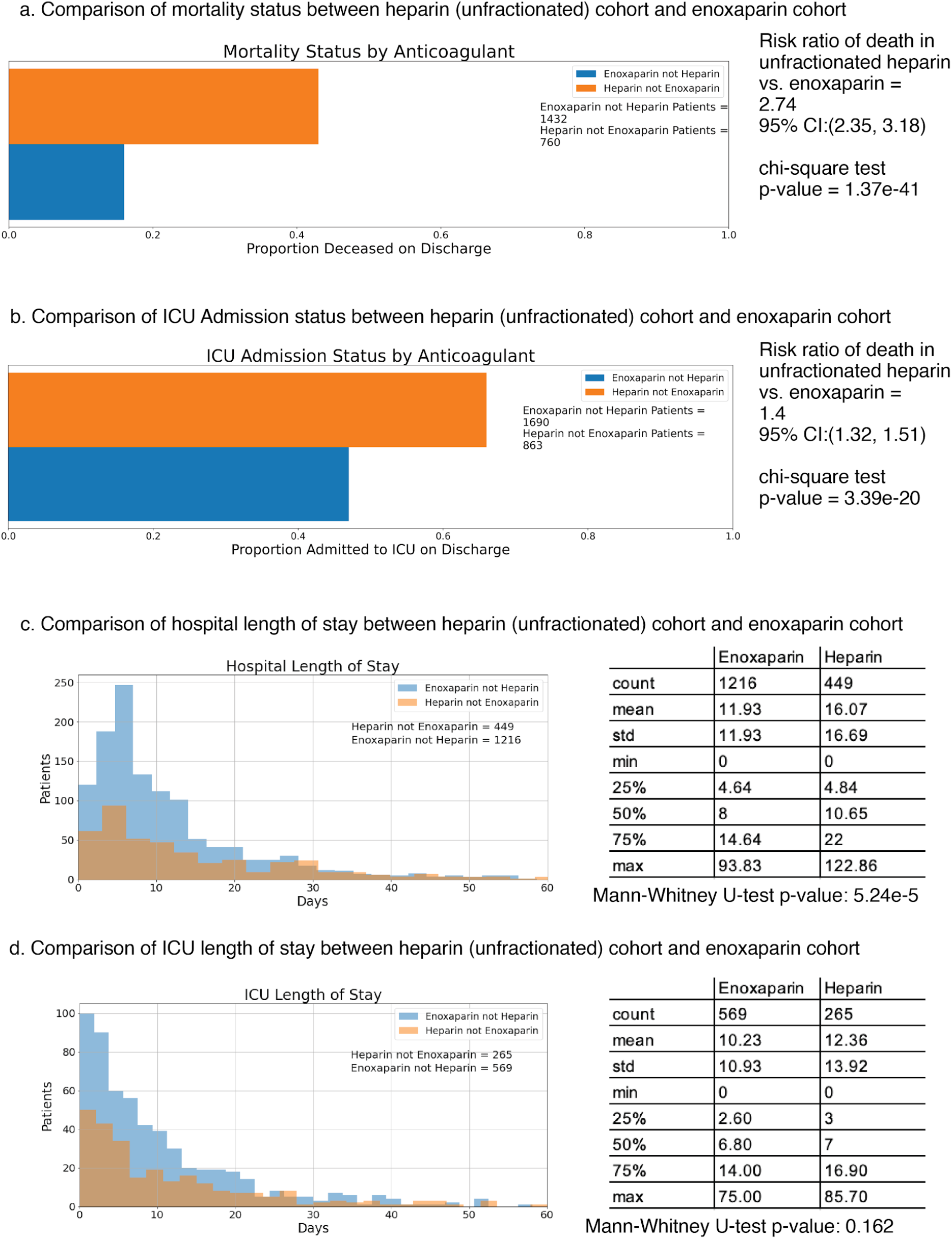
Comparison of outcomes between unfractionated heparin and enoxaparin patient cohorts (unadjusted). **(a)** Bar charts show a comparison of mortality status at discharge from the hospital between patient cohorts receiving enoxaparin but not heparin (**blue**) or unfractionated heparin but not enoxaparin (**orange**) during hospitalization **(b)** Bar charts show a comparison of mortality status at discharge from the hospital between patient cohorts receiving enoxaparin but not unfractionated heparin (**blue**) or unfractionated heparin but not enoxaparin (**orange**) during hospitalization **(c)** Histograms show ICU Length of Stay in days for cohorts of alive patients who received enoxaparin but not unfractionated heparin (**blue**) and reported a length of stay in the ICU and alive patients who received unfractionated heparin but not enoxaparin (**orange**) and reported a length of stay in the ICU **(d)** Histograms show hospital Length of Stay in days for cohorts of alive patients who received enoxaparin but not unfractionated heparin (**blue**) and reported a length of stay in the ICU and alive patients who received unfractionated heparin but not enoxaparin (**orange**) and reported a length of stay in the hospital.

Next, we compared the average lengths of stay in the ICU and hospital for the two cohorts. Here, we restricted the analysis to only patients that were alive at discharge. The length of stay in the ICU and in the hospital were shorter for the enoxaparin patients (mean ICU duration: 10.9 days, mean hospital duration: 11.9 days) compared to the unfractionated heparin patients (mean ICU duration: 13.9 days, mean hospital duration: 16.7 days) (**Figure 1c-d**). While the difference in average hospital length of stay is statistically significant (Mann-Whitney p-value: 5.2e-5), the difference in average ICU length of stay is not statistically significant (Mann-Whitney p-value: 0.16).

In **Figure 2**, we present the mortality rate and ICU admission rate for patients with different comorbidities: diabetes, hypertension, chronic kidney disease, and congestive heart failure. We observe that for the subgroups of patients with diabetes, hypertension, and congestive heart failure, patients administered enoxaparin have significantly lower rates of ICU admission and death compared to patients administered unfractionated heparin. For patients with chronic kidney disease, the difference in ICU admission rates between the enoxaparin and unfractionated heparin cohorts is statistically significant (risk ratio: 1.32, 95% C.I.: [1.06, 1.64], p-value: 0.01), however, the difference in mortality status is not statistically significant (risk ratio: 1.31, 95% C.I.: [0.94, 1.82], p-value: 0.12).

**Figure 2.**
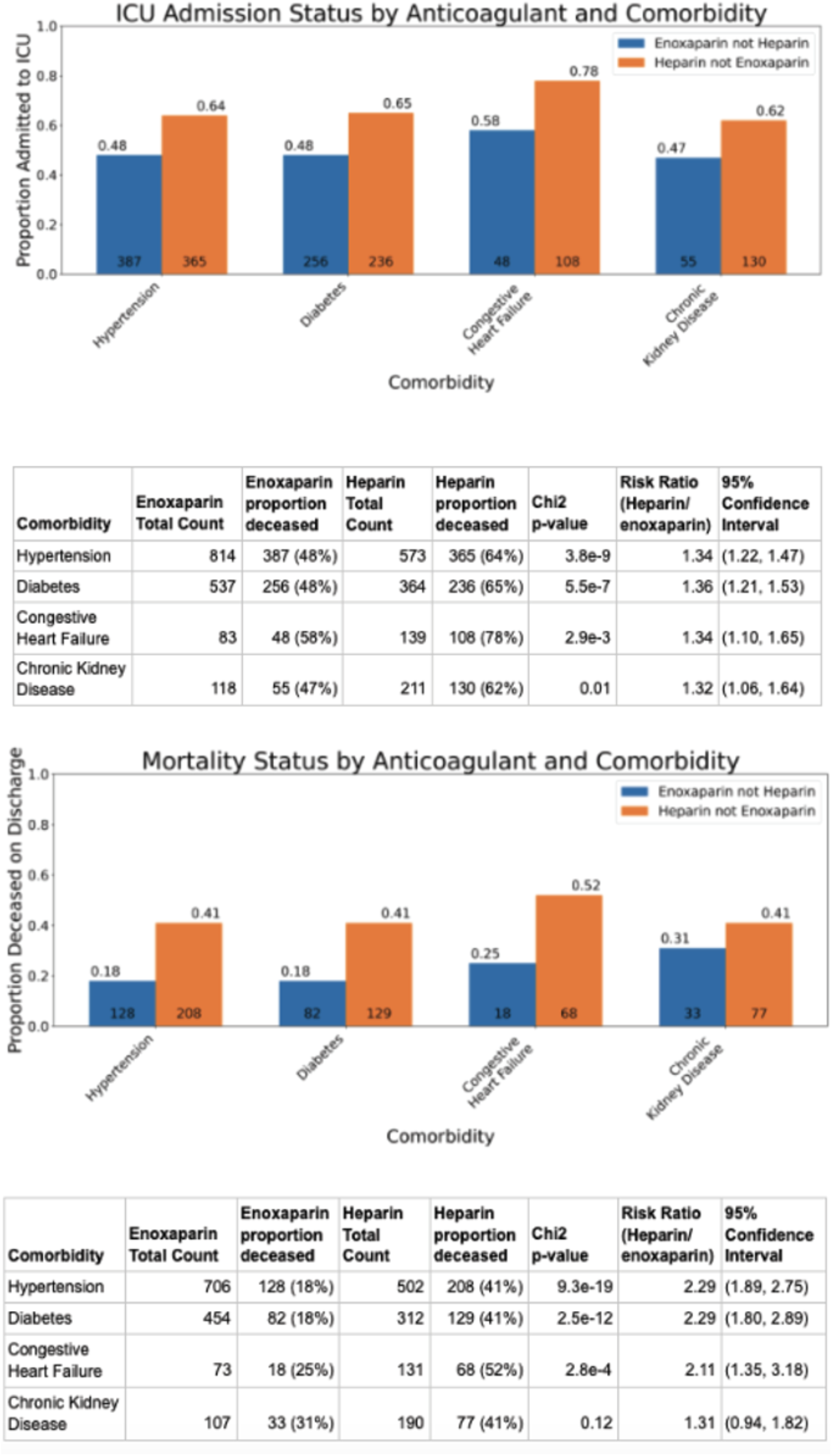
Comparison of outcomes between unfractionated heparin and enoxaparin patient cohorts in patients also reporting comorbidities. Bar charts show a comparison of Mortality Status at discharge from the hospital and status of admission to the ICU for 2 cohorts - patients receiving enoxaparin and reporting a comorbidity of interest (**blue**), and patients receiving heparin and reporting a comorbidity of interest (**orange**). Comorbidities include - Hypertension, Diabetes, Chronic Kidney Disease and Congestive Heart Failure. Statistics for these plots are included in the corresponding tables.

Next, we perform propensity score matching to control for a wide array of confounding factors simultaneously. The clinical characteristics of the matched and original unfractionated heparin and enoxaparin cohorts are shown in **Table 1**. Most covariates (including demographics, comorbidities, and admission diagnoses) appear well-matched. Of the 659 patients in the matched enoxaparin cohort, mortality status at discharge was available for 522, of which 119 (23%) were deceased on discharge; in the matched heparin cohort, information was available for 555 patients of which 215 (39%) were deceased on discharge (**Table 2**). This difference in mortality rates upon discharge was statistically significant (risk ratio: 1.70, 95% CI: [1.40, 2.05], adjusted p-value: 2.5e-7). The mortality rates reported at 28-days for both cohorts were consistent with the mortality rates reported upon hospital discharge, and differences in rates between the two cohorts were similarly statistically significant. Differences between the two cohorts in the average hospital and ICU length of stays were not statistically significant after matching.

**Table 1:**
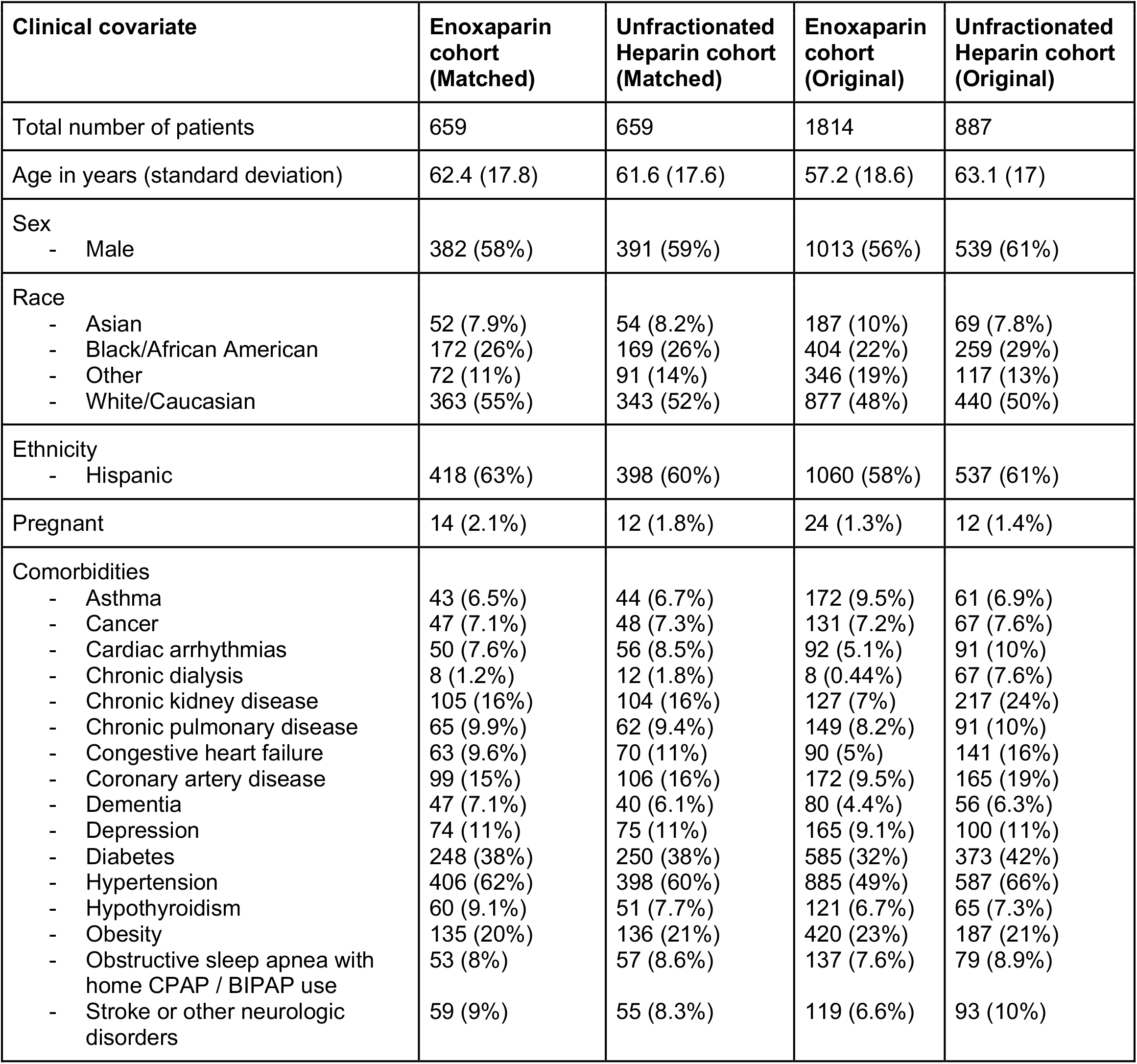
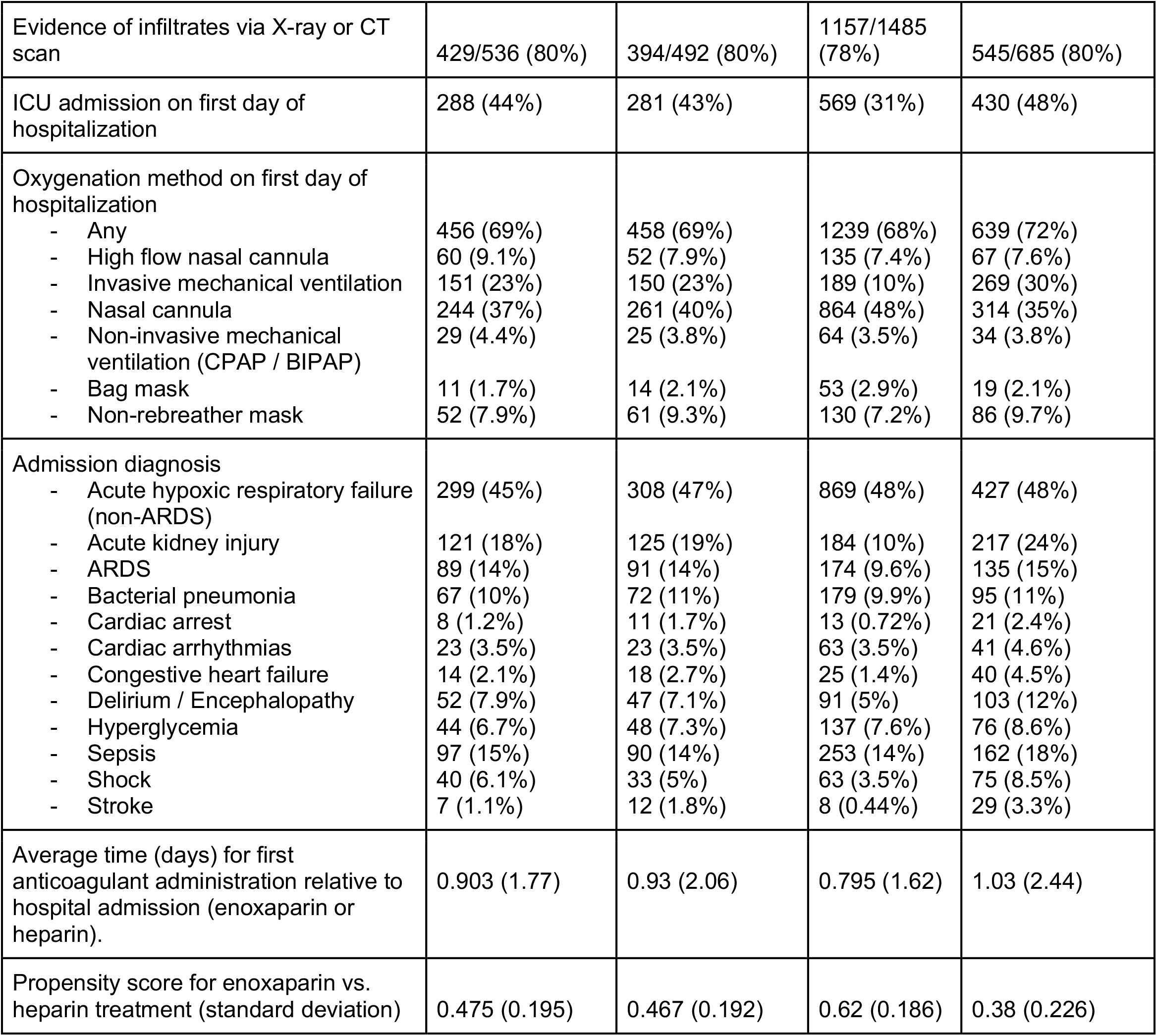
Covariate balancing results for Enoxaparin and Unfractionated Heparin cohorts. Summary of patient characteristics for matched and original cohorts of hospitalized COVID-19 patients who have taken either: unfractionated heparin or enoxaparin (but not both). For numeric variables such as age and first date of anticoagulant administration, the mean value for each cohort is shown with standard deviation in parentheses. For categorical variables such as race and ethnicity, patient counts are shown with the percentage of each cohort in parentheses. Denominators are shown when the variable has substantial missing data.

**Table 2:** Comparison of patient outcomes for Enoxaparin and Unfractionated Heparin cohorts. Summary of clinical outcomes for matched cohorts of hospitalized COVID-19 patients who have taken either unfractionated heparin or enoxaparin (but not both). For categorical variables such as mortality status and complications, patient counts are shown with the percentage of each cohort in parentheses. Only patients with reported outcomes in each cohort are used to determine the percentages. For numeric variables such as hospital and ICU length of stay, the mean value for each cohort is shown with standard deviation in parentheses. In addition, Benjamini-Hochberg adjusted p-values are shown for the statistical tests comparing the outcome variables for the matched enoxaparin and Heparin cohorts; relative risk of outcomes (heparin relative to enoxaparin) are shown as well, along with 95% confidence interval.

| Outcome variable | Enoxaparin cohort (Matched) (n=659) | Heparin cohort (Matched) (n=659) | BH-adjusted p-value | Relative risk (95% CI) Heparin vs. Enoxaparin |
| --- | --- | --- | --- | --- |
| Number of patients with reported outcomes |  |  |  |  |
| - Mortality status at hospital discharge (alive or deceased) | 522 | 555 |  |  |
| - Mortality status at 28 days (alive or deceased) | 522 | 555 |  |  |
| - ICU admission | 608 | 642 |  |  |
| - Hospital length of stay | 401 | 340 |  |  |
| - ICU length of stay | 223 | 189 |  |  |
| - Complications during hospitalization | 608 | 642 |  |  |
| Mortality at hospital discharge | 119/522 (23%) | 215/555 (39%) | 2.5e-07 | 1.70 (1.40, 2.05) |
| Mortality at 28 days | 127/522 (24%) | 227/555 (41%) | 2.3e-07 | 1.68 (1.40, 2.01) |
| ICU admission during hospitalization | 340/608 (56%) | 402/642 (63%) | 0.06 | 1.12 (1.02, 1.23) |
| Hospital length of stay (days) | 14 (13.8) | 14.2 (14.8) | 0.62 |  |
| ICU length of stay (days) | 11.2 (11.8) | 11.6 (12.7) | 0.95 |  |
| Complications during hospitalization |  |  |  |  |
| - Acute cardiac injury | 15 (2.5%) | 40 (6.2%) | 0.01 | 2.53 (1.39, 4.40) |
| - Acute kidney injury | 156 (26%) | 227 (35%) | 0.0019 | 1.38 (1.16, 1.63) |
| - ARDS | 152 (25%) | 184 (29%) | 0.36 | 1.15 (0.95, 1.38) |
| - Anemia | 46 (7.6%) | 81 (13%) | 0.02 | 1.67 (1.18, 2.34) |
| - Bacteremia | 24 (3.9%) | 32 (5%) | 0.62 | 1.26 (0.75, 2.10) |
| - Bacterial pneumonia | 65 (11%) | 69 (11%) | 0.95 | 1.01 (0.73, 1.38) |
| - Cardiac arrest | 48 (7.9%) | 62 (9.7%) | 0.58 | 1.22 (0.85, 1.75) |
| - Cardiac arrhythmia | 41 (6.7%) | 43 (6.7%) | 0.95 | 0.99 (0.66, 1.50) |
| - Co- or secondary infection | 34 (5.6%) | 49 (7.6%) | 0.36 | 1.36 (0.89, 2.07) |
| - Congestive heart failure | 17 (2.8%) | 21 (3.3%) | 0.91 | 1.17 (0.63, 2.17) |
| - Deep vein thrombosis | 17 (2.8%) | 20 (3.1%) | 0.95 | 1.11 (0.59, 2.08) |
| - Hyperglycemia | 65 (11%) | 75 (12%) | 0.83 | 1.09 (0.80, 1.49) |
| - Liver dysfunction | 34 (5.6%) | 44 (6.9%) | 0.62 | 1.23 (0.79, 1.88) |
| - Pleural Effusions | 15 (2.5%) | 27 (4.2%) | 0.30 | 1.70 (0.91, 3.10) |
| - Septic shock | 73 (12%) | 118 (18%) | 0.01 | 1.53 (1.17, 2.00) |
| - Stroke / Cerebrovascular incident | 5 (0.82%) | 17 (2.6%) | 0.07 | 3.22 (1.16, 7.81) |
| - Viral pneumonitis | 80 (13%) | 96 (15%) | 0.62 | 1.14 (0.86, 1.49) |

Information on complications that occurred after hospitalization was available for 608 of 659 patients in the matched enoxaparin cohort, and for 642 of 659 patients in the matched heparin cohort. Complications that occurred at a significantly higher rate in the matched heparin cohort compared to the matched enoxaparin cohort include: acute kidney injury (227 of 642 [35%] vs. 156 of 608 [26%] respectively, adjusted p-value 0.0019), acute cardiac injury (40 of 642 [6.2%] vs. 15 of 608 [2.5%] respectively, adjusted p-value 0.01), septic shock (118 of 642 [18%] vs. 73 of 608 [12%] respectively, adjusted p-value 0.01), and anemia (81 of 642 [13%] vs. 46 of 608 [7.6%,] respectively, adjusted p-value 0.02) (**Table 2**).

We also conducted an equivalent analysis comparing Enoxaparin vs other types of low molecular weight heparins (LMWH). The matching table is shown in **Table 3**. Of the 717 patients in this matched enoxaparin cohort, mortality status at discharge was available for 569 patients, of which 122 (21%) were deceased on discharge. In the 717 patients in the matched other LMWH cohort, information was available for 648 patients, of which 217 (33%) were deceased on discharge (risk ratio: 1.56, 95% C.I.: [1.29, 1.89]; adjusted p-value 4.4e-5) (**Table 4**). Complications which show statistically significant differences between the cohorts are ARDS (178 of 666 [27%] for enoxaparin vs. 236 of 697 [34%] for low molecular weight heparin; adjusted p-value: 0.03) and viral pneumonitis (125 of 66 [19%] for enoxaparin vs. 51 of 697 [7.3%] for low molecular weight heparin, adjusted p-value: 1.1e-8).

**Table 3.** Covariate balancing results for Enoxaparin and other LMWH cohorts. Summary of patient characteristics for matched and original cohorts of hospitalized COVID-19 patients who have taken either enoxaparin or some other low molecular weight Heparin (LMWH). For numeric variables such as age and first date of anticoagulant administration, the mean value for each cohort is shown with standard deviation in parentheses. For categorical variables such as race and ethnicity, patient counts are shown with the percentage of each cohort in parentheses. Denominators are shown for the covariates which have some missing data.

| Clinical covariate | Enoxaparin cohort (Matched) | Other LMWH cohort (Matched) | Enoxaparin cohort (Original) | LMWH cohort (Original) |
| --- | --- | --- | --- | --- |
| Total number of patients | 717 | 717 | 2045 | 857 |
| Age in years (standard deviation) | 59.9 (18.3) | 59.8 (18) | 57.6 (18.5) | 61.2 (17.6) |
| Sex |  |  |  |  |
| - Male | 397 (55%) | 429 (60%) | 1177 (58%) | 503 (59%) |
| Race |  |  |  |  |
| - Asian | 55 (7.7%) | 56 (7.8%) | 207 (10%) | 57 (6.7%) |
| - Black/African American | 166 (23%) | 166 (23%) | 489 (24%) | 189 (22%) |
| - Other | 111 (15%) | 129 (18%) | 388 (19%) | 155 (18%) |
| - White/Caucasian | 385 (54%) | 364 (51%) | 960 (47%) | 452 (53%) |
| Ethnicity |  |  |  |  |
| - Hispanic | 366 (51%) | 354/711 (50%) | 1214/2044 (59%) | 394/848 (46%) |
| Pregnant | 11 (1.5%) | 10 (1.4%) | 27 (1.3%) | 10 (1.2%) |
| Comorbidities |  |  |  |  |
| - Asthma | 53 (7.4%) | 53 (7.4%) | 204 (10%) | 56 (6.5%) |
| - Cancer | 55 (7.7%) | 57 (7.9%) | 148 (7.2%) | 67 (7.8%) |
| - Cardiac arrhythmias | 48 (6.7%) | 47 (6.6%) | 108 (5.3%) | 69 (8.1%) |
| - Chronic dialysis | 11 (1.5%) | 16 (2.2%) | 13 (0.64%) | 28 (3.3%) |
| - Chronic kidney disease | 85 (12%) | 71 (9.9%) | 161 (7.9%) | 99 (12%) |
| - Chronic pulmonary disease | 53 (7.4%) | 45 (6.3%) | 175 (8.6%) | 55 (6.4%) |
| - Congestive heart failure | 77 (11%) | 67 (9.3%) | 119 (5.8%) | 121 (14%) |
| - Coronary artery disease | 86 (12%) | 85 (12%) | 210 (10%) | 120 (14%) |
| - Dementia | 48 (6.7%) | 36 (5%) | 102 (5%) | 41 (4.8%) |
| - Depression | 59 (8.2%) | 54 (7.5%) | 199 (9.7%) | 63 (7.4%) |
| - Diabetes | 230 (32%) | 230 (32%) | 691 (34%) | 268 (31%) |
| - Hypertension | 388 (54%) | 357 (50%) | 1040 (51%) | 449 (52%) |
| - Hypothyroidism | 56 (7.8%) | 58 (8.1%) | 144 (7%) | 67 (7.8%) |
| - Obesity | 132 (18%) | 132 (18%) | 482 (24%) | 145 (17%) |
| - Obstructive sleep apnea with home CPAP / BIPAP use | 44 (6.1%) | 42 (5.9%) | 158 (7.7%) | 45 (5.3%) |
| - Stroke or other neurologic disorders | 64 (8.9%) | 58 (8.1%) | 138 (6.7%) | 78 (9.1%) |
| Evidence of infiltrates via X-ray or CT scan | 473/582 (81%) | 333/417 (80%) | 1355/1712 (79%) | 382/470 (81%) |
| ICU admission on first day of hospitalization | 316 (44%) | 311 (43%) | 650 (32%) | 412 (48%) |
| Oxygenation method on first day of hospitalization |  |  |  |  |
| - Any | 519 (72%) | 520 (73%) | 1422 (70%) | 635 (74%) |
| - High flow nasal cannula | 93 (13%) | 84 (12%) | 150 (7.3%) | 115 (13%) |
| - Invasive mechanical ventilation | 141 (20%) | 141 (20%) | 241 (12%) | 194 (23%) |
| - Nasal cannula | 266 (37%) | 276 (38%) | 1000 (49%) | 302 (35%) |
| - Non-invasive mechanical ventilation (CPAP / BIPAP) | 39 (5.4%) | 34 (4.7%) | 73 (3.6%) | 40 (4.7%) |
| - Bag mask | 30 (4.2%) | 34 (4.7%) | 52 (2.5%) | 44 (5.1%) |
| - Non-rebreather mask | 79 (11%) | 78 (11%) | 161 (7.9%) | 96 (11%) |
| Admission diagnosis |  |  |  |  |
| - Acute hypoxic respiratory failure (non-ARDS) | 352 (49%) | 353 (49%) | 1030 (50%) | 425 (50%) |
| - Acute kidney injury | 99 (14%) | 80 (11%) | 248 (12%) | 91 (11%) |
| - ARDS | 130 (18%) | 124 (17%) | 194 (9.5%) | 171 (20%) |
| - Bacterial pneumonia | 79 (11%) | 79 (11%) | 207 (10%) | 86 (10%) |
| - Cardiac arrest | 7 (0.98%) | 4 (0.56%) | 16 (0.78%) | 5 (0.58%) |
| - Cardiac arrhythmias | 32 (4.5%) | 21 (2.9%) | 68 (3.3%) | 29 (3.4%) |
| - Congestive heart failure | 22 (3.1%) | 17 (2.4%) | 29 (1.4%) | 29 (3.4%) |
| - Delirium / Encephalopathy | 55 (7.7%) | 43 (6%) | 113 (5.5%) | 59 (6.9%) |
| - Hyperglycemia | 51 (7.1%) | 45 (6.3%) | 170 (8.3%) | 46 (5.4%) |
| - Sepsis | 103 (14%) | 96 (13%) | 328 (16%) | 107 (12%) |
| - Shock | 34 (4.7%) | 30 (4.2%) | 88 (4.3%) | 32 (3.7%) |
| - Stroke | 4 (0.56%) | 6 (0.84%) | 8 (0.39%) | 23 (2.7%) |
| Day of first anticoagulant administration relative to hospital admission (enoxaparin or heparin) | 0.997 (2.16) | 0.907 (2.2) | 0.973 (1.99) | 0.847 (2.08) |
| Propensity score for enoxaparin vs. low-molecular-weight heparin treatment (standard deviation) | 0.468 (0.178) | 0.462 (0.175) | 0.582 (0.166) | 0.418 (0.193) |

**Table 4:** Comparison of patient outcomes for Enoxaparin and other LMWH cohorts. Summary of clinical outcomes for matched cohorts of hospitalized COVID-19 patients who have taken either enoxaparin or other low-molecular-weight heparins (LMWH). For categorical variables such as mortality status and complications, patient counts are shown with the percentage of each cohort in parentheses. Only patients with reported outcomes in each cohort are used to determine the percentages. For numeric variables such as hospital and ICU length of stay, the mean value for each cohort is shown with standard deviation in parentheses. In addition, Benjamini-Hochberg adjusted p-values are shown for the statistical tests comparing the outcome variables for the matched enoxaparin and Heparin cohorts.

| Outcome variable | Enoxaparin cohort (Matched) (n=717) | Other LMWH cohort (Matched) (n=717) | BH-adjusted p-value | Relative risk (95% CI), other LMWH vs. Enoxaparin |
| --- | --- | --- | --- | --- |
| Number of patients with reported outcomes |  |  |  |  |
| - Mortality status at hospital discharge (alive or deceased) | 569 | 648 |  |  |
| - Mortality status at 28 days (alive or deceased) | 569 | 648 |  |  |
| - ICU admission | 666 | 697 |  |  |
| - Hospital length of stay | 446 | 429 |  |  |
| - ICU length of stay | 273 | 257 |  |  |
| - Complications during hospitalization | 666 | 697 |  |  |
| Mortality at hospital discharge | 122/569 (21%) | 217/648 (33%) | 4.4e-05 | 1.56 (1.29, 1.89) |
| Mortality at 28 days | 130/569 (23%) | 223/648 (34%) | 9e-05 | 1.51 (1.25, 1.81) |
| ICU admission during hospitalization | 400/666 (60%) | 460 (66%) | 0.08 | 1.10 (1.01, 1.19) |
| Hospital length of stay (days) | 14.8 (13.2) | 15.8 (12.9) | 0.09 |  |
| ICU length of stay (days) | 10.8 (10.2) | 10.4 (9.83) | 0.96 |  |
| Complications during hospitalization |  |  |  |  |
| - Acute cardiac injury | 21 (3.2%) | 42 (6%) | 0.07 | 1.91 (1.14, 3.14) |
| - Acute kidney injury | 150 (23%) | 139 (20%) | 0.43 | 0.89 (0.72, 1.09) |
| - ARDS | 178 (27%) | 236 (34%) | 0.03 | 1.27 (1.08, 1.49) |
| - Anemia | 85 (13%) | 72 (10%) | 0.32 | 0.81 (0.60, 1.09) |
| - Bacteremia | 40 (6%) | 28 (4%) | 0.22 | 0.67 (0.42, 1.07) |
| - Bacterial pneumonia | 83 (12%) | 60 (8.6%) | 0.08 | 0.69 (0.51, 0.95) |
| - Cardiac arrest | 56 (8.4%) | 64 (9.2%) | 0.79 | 1.09 (0.78, 1.53) |
| - Cardiac arrhythmia | 42 (6.3%) | 29 (4.2%) | 0.21 | 0.66 (0.42, 1.05) |
| - Co- or secondary infection | 44 (6.6%) | 43 (6.2%) | 0.87 | 0.93 (0.62, 1.40) |
| - Congestive heart failure | 18 (2.7%) | 12 (1.7%) | 0.43 | 0.64 (0.32, 1.31) |
| - Deep vein thrombosis | 22 (3.3%) | 17 (2.4%) | 0.59 | 0.74 (0.40, 1.38) |
| - Hyperglycemia | 83 (12%) | 67 (9.6%) | 0.22 | 0.77 (0.57, 1.05) |
| - Liver dysfunction | 60 (9%) | 41 (5.9%) | 0.09 | 0.65 (0.45, 0.96) |
| - Pleural Effusions | 23 (3.5%) | 21 (3%) | 0.83 | 0.87 (0.49, 1.55) |
| - Septic shock | 98 (15%) | 96 (14%) | 0.79 | 0.94 (0.72, 1.21) |
| - Stroke / Cerebrovascular incident | 9 (1.4%) | 13 (1.9%) | 0.76 | 1.38 (0.60, 3.09) |
| - Viral pneumonitis | 125 (19%) | 51 (7.3%) | 1.1e-08 | 0.39 (0.29, 0.53) |

We also examined whether there were any race-based differences in the administration of enoxaparin and unfractionated heparin. The cohorts of interest were Black/African American patients with anticoagulant information available (n = 1,203) and White/Caucasian patients with anticoagulant information available (n = 2,488). Propensity score matching was performed, and the clinical characteristics of the matched and original cohorts are shown in **Table 5**. We observe that the clinical covariates are well-balanced for the matched cohorts. Rates of administration of unfractionated heparin, enoxaparin, and other LMWH medications for the original and matched cohorts are shown in **Tables 6** and **7**, respectively.

**Table 5.**
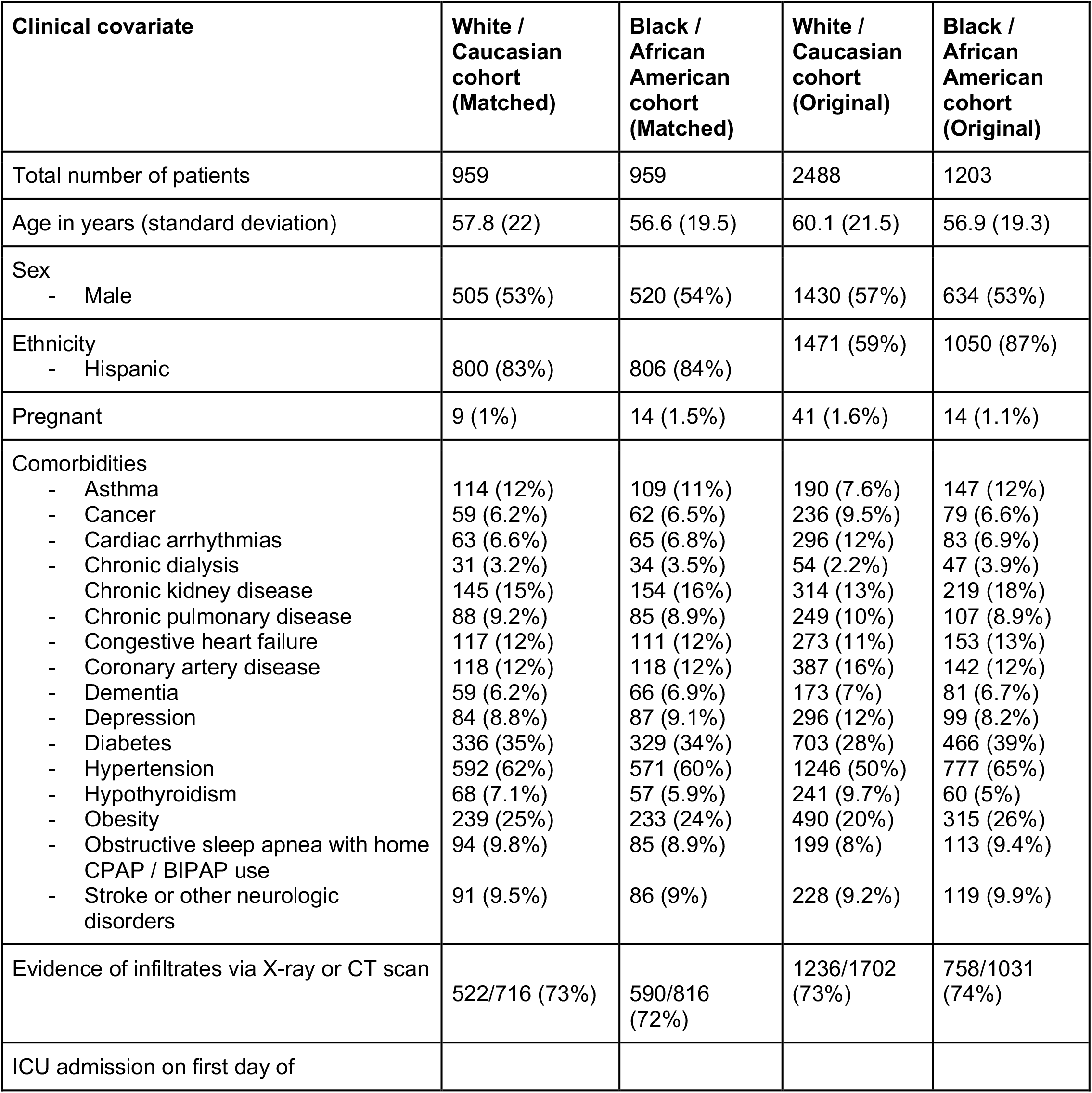
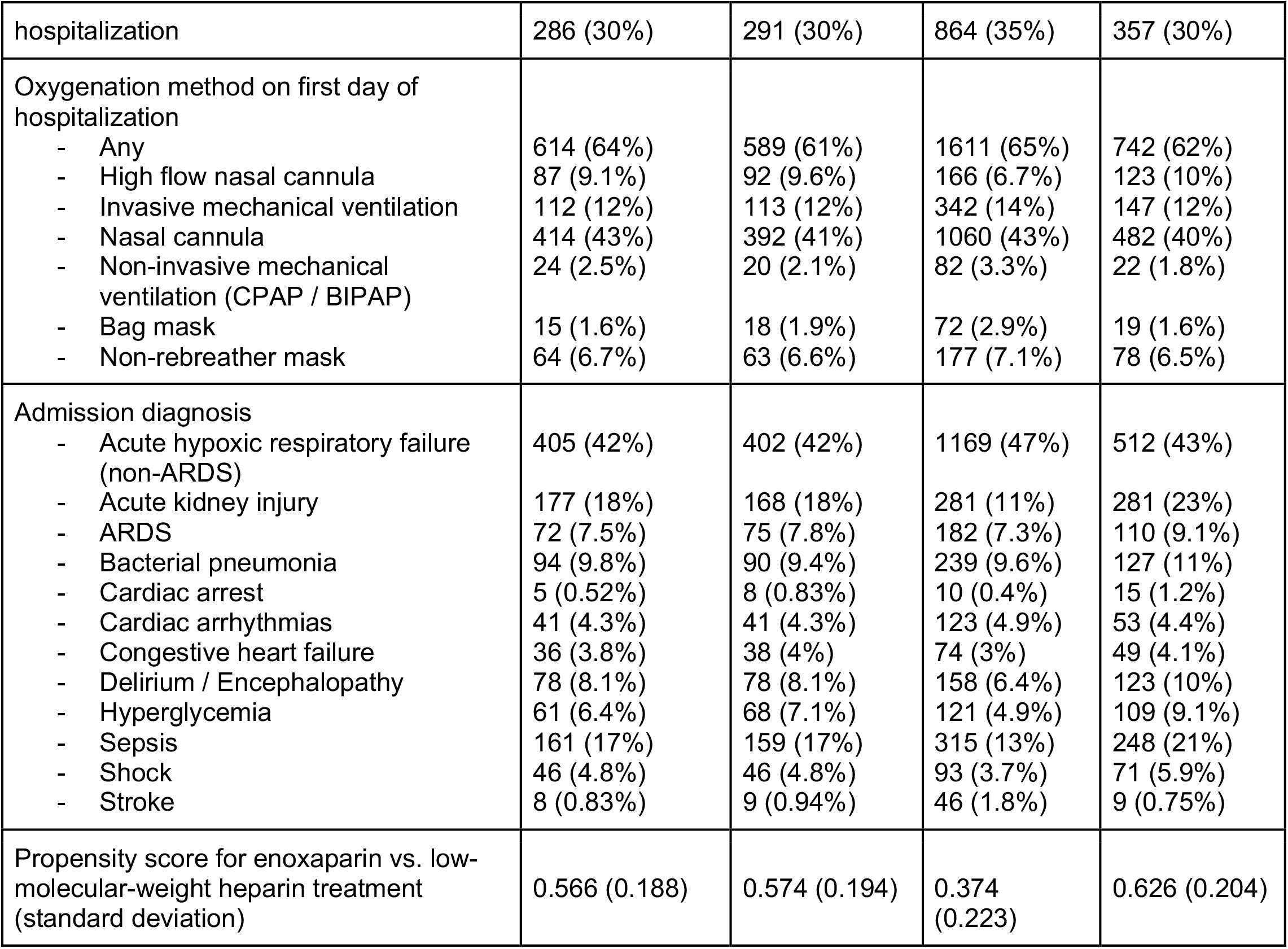
Covariate balancing results for race-based cohorts. Summary of patient characteristics for matched and original cohorts of Black/African American and White/Caucasian hospitalized COVID-19 patients. For numeric variables such as age and first date of anticoagulant administration, the mean value for each cohort is shown with standard deviation in parentheses. For categorical variables such as race and ethnicity, patient counts are shown with the percentage of each cohort in parentheses.

| **Clinical covariate** | **White /**<br>**Caucasian**<br>**cohort**<br>**(Matched)** | **Black /**<br>**African**<br>**American**<br>**cohort**<br>**(Matched)** | **White /**<br>**Caucasian**<br>**cohort**<br>**(Original)** | **Black /**<br>**African**<br>**American**<br>**cohort**<br>**(Original)** |
| --- | --- | --- | --- | --- |
| Total number of patients | 959 | 959 | 2488 | 1203 |
| Age in years (standard deviation) | 57.8 (22) | 56.6 (19.5) | 60.1 (21.5) | 56.9 (19.3) |
| Sex<br>- Male | 505 (53%) | 520 (54%) | 1430 (57%) | 634 (53%) |
| Ethnicity<br>- Hispanic | 800 (83%) | 806 (84%) | 1471 (59%) | 1050 (87%) |
| Pregnant | 9 (1%) | 14 (1.5%) | 41 (1.6%) | 14 (1.1%) |
| Comorbidities<br>- Asthma<br>- Cancer<br>- Cardiac arrhythmias<br>- Chronic dialysis<br>- Chronic kidney disease<br>- Chronic pulmonary disease<br>- Congestive heart failure<br>- Coronary artery disease<br>- Dementia<br>- Depression<br>- Diabetes<br>- Hypertension<br>- Hypothyroidism<br>- Obesity<br>- Obstructive sleep apnea with home CPAP / BIPAP use<br>- Stroke or other neurologic disorders | 114 (12%)<br>59 (6.2%)<br>63 (6.6%)<br>31 (3.2%)<br>145 (15%)<br>88 (9.2%)<br>117 (12%)<br>118 (12%)<br>59 (6.2%)<br>84 (8.8%)<br>336 (35%)<br>592 (62%)<br>68 (7.1%)<br>239 (25%)<br>94 (9.8%)<br>91 (9.5%) | 109 (11%)<br>62 (6.5%)<br>65 (6.8%)<br>34 (3.5%)<br>154 (16%)<br>85 (8.9%)<br>111 (12%)<br>118 (12%)<br>66 (6.9%)<br>87 (9.1%)<br>329 (34%)<br>571 (60%)<br>57 (5.9%)<br>233 (24%)<br>85 (8.9%)<br>86 (9%) | 190 (7.6%)<br>236 (9.5%)<br>296 (12%)<br>54 (2.2%)<br>314 (13%)<br>249 (10%)<br>273 (11%)<br>387 (16%)<br>173 (7%)<br>296 (12%)<br>703 (28%)<br>1246 (50%)<br>241 (9.7%)<br>490 (20%)<br>199 (8%)<br>228 (9.2%) | 147 (12%)<br>79 (6.6%)<br>83 (6.9%)<br>47 (3.9%)<br>219 (18%)<br>107 (8.9%)<br>153 (13%)<br>142 (12%)<br>81 (6.7%)<br>99 (8.2%)<br>466 (39%)<br>777 (65%)<br>60 (5%)<br>315 (26%)<br>113 (9.4%)<br>119 (9.9%) |
| Evidence of infiltrates via X-ray or CT scan | 522/716 (73%) | 590/816 (72%) | 1236/1702 (73%) | 758/1031 (74%) |
| ICU admission on first day of |  |  |  |  |
| hospitalization | 286 (30%) | 291 (30%) | 864 (35%) | 357 (30%) |
| Oxygenation method on first day of hospitalization |  |  |  |  |
| - Any | 614 (64%) | 589 (61%) | 1611 (65%) | 742 (62%) |
| - High flow nasal cannula | 87 (9.1%) | 92 (9.6%) | 166 (6.7%) | 123 (10%) |
| - Invasive mechanical ventilation | 112 (12%) | 113 (12%) | 342 (14%) | 147 (12%) |
| - Nasal cannula | 414 (43%) | 392 (41%) | 1060 (43%) | 482 (40%) |
| - Non-invasive mechanical ventilation (CPAP / BIPAP) | 24 (2.5%) | 20 (2.1%) | 82 (3.3%) | 22 (1.8%) |
| - Bag mask | 15 (1.6%) | 18 (1.9%) | 72 (2.9%) | 19 (1.6%) |
| - Non-rebreather mask | 64 (6.7%) | 63 (6.6%) | 177 (7.1%) | 78 (6.5%) |
| Admission diagnosis |  |  |  |  |
| - Acute hypoxic respiratory failure (non-ARDS) | 405 (42%) | 402 (42%) | 1169 (47%) | 512 (43%) |
| - Acute kidney injury | 177 (18%) | 168 (18%) | 281 (11%) | 281 (23%) |
| - ARDS | 72 (7.5%) | 75 (7.8%) | 182 (7.3%) | 110 (9.1%) |
| - Bacterial pneumonia | 94 (9.8%) | 90 (9.4%) | 239 (9.6%) | 127 (11%) |
| - Cardiac arrest | 5 (0.52%) | 8 (0.83%) | 10 (0.4%) | 15 (1.2%) |
| - Cardiac arrhythmias | 41 (4.3%) | 41 (4.3%) | 123 (4.9%) | 53 (4.4%) |
| - Congestive heart failure | 36 (3.8%) | 38 (4%) | 74 (3%) | 49 (4.1%) |
| - Delirium / Encephalopathy | 78 (8.1%) | 78 (8.1%) | 158 (6.4%) | 123 (10%) |
| - Hyperglycemia | 61 (6.4%) | 68 (7.1%) | 121 (4.9%) | 109 (9.1%) |
| - Sepsis | 161 (17%) | 159 (17%) | 315 (13%) | 248 (21%) |
| - Shock | 46 (4.8%) | 46 (4.8%) | 93 (3.7%) | 71 (5.9%) |
| - Stroke | 8 (0.83%) | 9 (0.94%) | 46 (1.8%) | 9 (0.75%) |
| Propensity score for enoxaparin vs. low-molecular-weight heparin treatment (standard deviation) | 0.566 (0.188) | 0.574 (0.194) | 0.374 (0.223) | 0.626 (0.204) |

**Table 6.** Anticoagulant administration rates by race among unmatched cohorts. Rates of anticoagulant administration among unmatched Black/African American and White/Caucasian cohorts of hospitalized COVID-19 patients.

| **Anticoagulant administered** | **Black / African American cohort (n=1,203)** | **White / Caucasian cohort (n=2,488)** | **Chi-square test p-value** | **BH-corrected p-value** | **Relative risk (95% CI)** |
| --- | --- | --- | --- | --- | --- |
| Enoxaparin | 520 (43%) | 1032 (41%) | 0.33 | 0.34 | 1.04 (0.96, 1.13) |
| Unfractionated heparin | 375 (31%) | 595 (24%) | 3.2E-06 | 1.6E-05 | 1.30 (1.17, 1.45) |
| Other low molecular weight heparin | 220 (18%) | 524 (21%) | 0.05 | 0.08 | 0.87 (0.75, 1.00) |
| Enoxaparin but not unfractionated heparin | 404 (34%) | 877 (35%) | 0.34 | 0.34 | 0.95 (0.87, 1.05) |
| Unfractionated heparin but not enoxaparin | 259 (22%) | 440 (18%) | 0.006 | 0.015 | 1.22 (1.06, 1.40) |

Looking at the original unmatched cohorts, Black/African American patients had significantly higher rates of administration of unfractionated heparin compared to White/Caucasian patients (375 of 1203 [31%] vs. 595 of 2488 [24%] respectively, adjusted p-value: 1.6e-05) (**Table 6**). After matching, this difference in unfractionated heparin use remains statistically significant (272 of 959 [28%] for Black/African American patients vs. 213 of 959 [22%] for White/Caucasian patients, adjusted p-value: 0.01) (**Table 7**). On the other hand, enoxaparin and other low molecular weight heparins are administered at similar rates in the unmatched and matched cohorts (**Tables 6-7**). Finally, the proportion of patients which took exclusively either enoxaparin or unfractionated heparin are similar for the Black/African American and White/Caucasian cohorts.

**Table 7.** Anticoagulant administration rates by race among matched cohorts. Rates of anticoagulant administration among matched Black/African American and White/Caucasian cohorts of hospitalized COVID-19 patients.

| **Anticoagulant administered** | **Black / African American cohort (Matched) (n=959)** | **White / Caucasian cohort (Matched) (n=959)** | **Chi-square test p-value** | **BH-corrected p-value** | **Relative risk (95% CI)** |
| --- | --- | --- | --- | --- | --- |
| Enoxaparin | 418 (44%) | 419 (44%) | 1 | 1 | 1.00 (0.9, 1.10) |
| Unfractionated heparin | 272 (28%) | 213 (22%) | 0.002 | 0.01 | 1.28 (1.09, 1.49) |
| Other low molecular weight heparin | 186 (19%) | 186 (19%) | 1 | 1 | 1.00 (0.83, 1.20) |
| Enoxaparin but not unfractionated heparin | 332 (35%) | 362 (38%) | 0.17 | 0.28 | 0.91 (0.81, 1.03) |
| Unfractionated heparin but not enoxaparin | 186 (19%) | 156 (16%) | 0.08 | 0.2 | 1.19 (0.98, 1.44) |

## Discussion

Prior work has shown that anticoagulant treatments and prophylaxis are associated with improved outcomes for COVID-19 patients^9,10,^. In particular, there is evidence to suggest that low molecular weight heparin can be used to effectively treat COVID-19 patients with coagulopathy^11^. This retrospective analysis suggests that enoxaparin, a particular form of low molecular weight heparin, shows promise as an anticoagulant therapy for severe COVID-19, compared to both unfractionated heparin and other low molecular weight heparin therapies. These findings are consistent with a retrospective study on electronic health records from the Mayo Clinic which has found that enoxaparin is associated with lower rates of thrombotic events, kidney injury, and mortality in comparison with unfractionated heparin^12^. However, this study goes beyond the previous analysis by leveraging the massive SCCM VIRUS data registry of hospitalized COVID-19 patients from multiple sites around the world. As a result, we find that there are additional complications which are enriched at a statistically significant level in the unfractionated heparin cohort compared to the enoxaparin cohort, including septic shock and anemia.

There are several limitations of this study. While we have longitudinal data from the registry on daily anticoagulant use, we do not have access to the detailed physician notes for these patients. Therefore, in this dataset we cannot distinguish between prophylactic and therapeutic anticoagulant use. Since we include only patients who received an anticoagulant medication, there is potential for immortal time bias because there may be some patients who died prior to anticoagulant administration in the hospital. Another limitation of this study is the lack of follow-up data for all patients. For many sites, we do not have access to follow-up data for patients to determine 28-day mortality status, so the mortality rates may be skewed towards the sites of the study where this outcome data is most available. Finally, there are differences in the FDA drug labels for unfractionated heparin, enoxaparin, and other forms of low molecular weight heparin, which can lead to differences in real-world patterns of prescription^13,14^. For example, patients with active kidney disease are contraindicated for higher doses of enoxaparin. However, unfractionated heparin does not require any dose modifications for patients with active kidney disease, so there may be a preference for this medication among this cohort of patients. Although these biases in prescription patterns are partially controlled for by the propensity score matching algorithm, there may be some additional unobserved confounding factors which are not taken into consideration.

There are numerous follow-up analyses which may be inspired from this study. As more data becomes available, we may investigate differential patient outcomes for other variants of low molecular weight heparin beyond enoxaparin. Similar comparative analyses may be undertaken for other COVID-19 treatment options beyond anticoagulants, such as supplemental oxygenation methods. This study demonstrates the utility of the SCCM VIRUS data registry for analyzing diverse research questions related to therapeutics for severe COVID-19 patients^5^. Finally, a number of studies have been analyzing the association between race/ethnicity and clinical outcomes in COVID-19^15,16^. From this study, the race-associated differences in the administration of the anticoagulants enoxaparin and unfractionated heparin warrants further analyses into the associations between patients’ race/ethnicity, comorbidities, and administration of medications in managing COVID-19.

## Data Availability

All reasonable requests for aggregated data should be routed to the corresponding author and will be referred to the VIRUS registry for further processing.

## Acknowledgements

We thank the SCCM Discovery VIRUS data registry and the Collaborative Co-authors listed in *Supplementary Table S4* for providing and maintaining the database of hospitalized COVID-19 patients which made this study possible. We are grateful to Vishakha Kumar for all the timely help and inputs, and to the SCCM colleagues for their internal review and feedback on this study. We are particularly thankful to Ognjen Gajic of the Mayo Clinic and Allan Walkey of Boston University for their helpful contributions to this study. Finally, we would like to thank Murali Aravamudan and Patrick Lenehan of nference for their reviews and feedback this manuscript.

## Supplementary Material

**Supplementary Table S1.** Frequency of administration of different anticoagulants.

| Anticoagulant | Patient Count |
| --- | --- |
| Enoxaparin | 2175 |
| Unfractionated Heparin | 1248 |
| Low Molecular Weight Heparin | 987 |
| Apixaban | 233 |
| Other | 210 |
| Rivaroxaban | 64 |
| Vitamin K antagonist | 34 |
| Edoxaban | 27 |
| Argatroban | 17 |
| Fondaparinux | 13 |
| Dabigatran | 12 |
| Bivalirudin | 8 |
| TPA/RTPA | 8 |

**Supplementary Table S2:** Diagnostic criteria for complications listed in the VIRUS registry questionnaire.

|  |  |
| --- | --- |
| Acute Kidney Injury | Must meet stage 1 AKI criteria or higher as defined by 2012 Kidney Disease: Improving Global Outcomes (KDIGO) Clinical Practice Guideline for Acute Kidney Injury (AKI)<br>Stage 1 Criteria:<br>1) serum Cr increases by 1.5-1.9 above baseline or $\geq 0.3$mg/dl increase in serum Cr.<br>2) Urine output $< 0.5$ ml/Kg/hour for 6-12 hours |
| Congestive Heart Failure | Heart Failure is a complex clinical syndrome resulting from any structural or functional cardiac disorder that impairs the ability of the ventricle to fill with or eject blood. It is characterized by dyspnea and fatigue in the medical history and edema, rales on physical examination. |
|  | New York Heart Association (NYHA) Classification - The Stages of Heart Failure:<br>Class I - No symptoms and no limitation in ordinary physical activity, e.g. shortness of breath when walking, climbing stairs etc.<br>Class II - Mild symptoms (mild shortness of breath and/or angina) and slight limitation during ordinary activity.<br>Class III - Marked limitation in activity due to symptoms, even during less-than-ordinary activity, e.g. walking short distances (20—100 m). Comfortable only at rest.<br>Class IV - Severe limitations. Experiences symptoms even while at rest. Mostly bed bound patients. |

**Supplementary Table S3.** Data completeness among the 5,065 patients with anticoagulant information available.

| **Covariate** | **Number of patients with data available** |
| --- | --- |
| Total Patients | 5065 |
| Demographics<br>- Age<br>- Sex<br>- Race<br>- Ethnicity (Hispanic/non-Hispanic) | 4993 (99%)<br>5065 (100%)<br>5051 (100%)<br>5047 (100%) |
| Comorbidities | 4681 (92%) |
| Admission diagnoses | 4985 (98%) |
| Oxygenation methods | 4949 (98%) |
| Evidence of infiltrates (X-ray or CT scan) | 4058 (80%) |

**Supplementary Table S4 – Collaborative Co-authors List:**

**<u>Argentina</u>**

**Hospital Universitario Austral:** Ana Julieta Herrera

**<u>Belgium</u>**

**Centre Hospitalier Jolimont:** Jean-Baptiste Mesland, Pierre Henin, Hélène Petre, Isabelle Buelens, Anne-Catherine Gerard

**The Brugmann University Hospital, Bruxelles:** Philippe Clevenbergh

**<u>Bolvia</u>**

**Clinica Los Olivos:** Rolando Claure-Del Granado, Jose A. Mercado, Esdenka Vega-Terrazas, Maria F. Iturricha-Caceres

**<u>Bosnia and Herzegovina</u>**

**University Clinical Hospital, Mostar:** Dragana Markotić, Ivana Bošnjak

**University Clinical Centre of the Republic of Srpska, Banja Luka:** Pedja Kovacevic

**<u>Columbia</u>**

**Clinica Medical SAS:** Oscar Y Gavidia, Felipe Pachon, Yeimy A Sanchez

**<u>Croatia</u>**

**Clinical Hospital Center Rijeka:** Danijel Knežević

**<u>Egypt</u>**

**Helwan University:** Mohamed El Kassas, Mohamed Badr, Ahmed Tawheed, Ahmed Tawheed, Hend Yahia

**<u>Hondurus</u>**

**CEMESA Hospital:** Sierra-Hoffman, Fernando Valerio, Oscar Diaz

**Honduras Medical Center:** Jose Luis Ramos Coello, Guillermo Perez, Ana Karen Vallecillo Lizardo, Gabina María Reyes Guillen, Helin Archaga Soto

**<u>Hungary</u>**

**Uzsoki Teaching Hospital:** Csaba Kopitkó, Ágnes Bencze, István Méhész, MD, Zsófia Gerendai

**<u>India</u>**

**ACSR Govt. Medical College and Hospital:** Neethi Chandra

**Jawaharlal Institute of Postgraduate Medical Education and Research:** Anusha Cherian, Sreejith Parameswaran, Magesh Parthiban, Menu Priya A.

**KLEs Dr. Prabhakar Kore Hospital & MRC:** Madhav Prabhu, Vishal Jakati

**Maulana Azad Medical College and Lok Nayak Hospital:** Mradul Kumar Daga, Munisha Agarwal, Ishan Rohtagi

**Rising Medicare Hospital:** Giri Deepak Ramgir

**<u>Japan</u>**

**Center Hospital of the National Center for Global Health and Medicine:** Wataru Matsuda, Reina Suzuki

**Hiroshima University:** Michihito Kyo

**Sapporo City General Hospital:** Yuki Itagaki, Akira Kodate, Reina Suzuki, Akira Kodate,Yuki Takahashi, Koyo Moriki

**Tokyo Medical and Dental University:** Hidenobu Shigemitsu, Yuka Mishima, Nobuyuki Nosaka, Michio Nagashima

**<u>Mexico</u>**

**Centenario Hospital Miguel Hidalgo:** Mariana Janeth Hermosillo Ulloa

**<u>Pakistan</u>**

**Nishtar Hospital Multan:** Muhammad H Khan, Muhammad Tayyeb

**The Aga Khan University Hospital:** Sidra Ishaque, Ali Faisal Saleem, Naved Rahman Siddiqui, Salima Sherali, Yasmin Hashwani, ShafiaI Shaque

**<u>Peru</u>**

**Cayetano Heredia National Hospital:** Juan Carrasco

**<u>Puerto Rico</u>**

**San Juan City Hospital:** Ricardo Alan Hernandez Castillo, Hector Omar Collazo Santiago

**Hospital Auxilio Mutuo:** Ricardo Alan Hernandez, Héctor Collazo Santiago, Héctor Collazo Santiago

**<u>Russia</u>**

**Kuban State Medical University with affiliation Territorial Hospital #2:** Igor Borisovich Zabolotskikh, Konstantin Dmitrievich Zybin, Sergey Vasilevich Sinkov, Tatiana Sergeevna Musaeva

**<u>Saudi Arabia</u>**

**King Fahad Armed Forces Hospital:** Razan K Alamoudi, Hassan M. AlSharif, Sarah A. Almazwaghi, Mohammed S Elsakran, Mohamed A Aid, Mouaz A Darwich, Omnia M Hagag, Salah A Ali, Alona Rocacorba, Kathrine Supeña, Efren Ray Juane, Jenalyn Medina, Jowany Baduria

**King Faisal Specialist Hospital & Research Centre:** Marwa Ridha Amer, Mohammed Abdullah Bawazeer, Talal I. Dahhan, Eiad Kseibi, Abid Shahzad Butt, Syed Moazzum Khurshid, Muath Rabee, Mohammed Abujazar, Razan Alghunaim, Maal Abualkhair, Abeer Turki AlFirm, Eiad Kseibi, Syed Moazzum Khurshid, Muath Rabee, Mohammed Abujazar, Razan Alghunaim

**King Saud bin Abdulaziz University for Health Sciences and King Abdullah International Medical Research Center:** Yaseen M Arabi, Sheryl Ann Abdukahil

**King Saud University:** Mohammed A Almazyad, Mohammed I Alarifi, Jara M Macarambon, Ahmad Abdullah Bukhari, Hussain A. Albahrani, Kazi N Asfina, Kaltham M Aldossary

**<u>Serbia</u>**

**Clinical Centre of Vojvodina, Novi Sad:** Gordana Jovanovic

**University Hospital Center “Dr Dragisa Misovic-Dedinje”:** Predrag D Stevanovic, Dejan S Stojakov, Duska K Ignjatovic, Suzana C Bojic, Marina M Bobos, Irina B Nenadic, Milica S Zaric, Marko D Djuric, Vladimir R Djukic

**<u>Spain</u>**

**Hospital Universitario La Paz:** Santiago Y. Teruel, Belen C. Martin,Santiago Y. Teruel

**Hospital Universitario, Universidad Autonoma de Nuevo León:** Rene Rodriguez-Gutierrez, Jose Gerardo Gonzalez-Gonzalez, Alejandro Salcido-Montenegro, Adrian Camacho-Ortiz

**<u>United States of America</u>**

**Advocate Children’s Hospital, IL:** Varsha P Gharpure, Usman Raheemi

**Advocate Christ Medical Center:** Kenneth W. Dodd, Nicholas Goodmanson, Kathleen Hesse,Paige Bird, Chauncey Weinert, Nathan Schoenrade, Abdulrahman Altaher, Esmael Mayar, Matthew Aronson, Tyler Cooper, Monica Logan, Brianna Miner, Gisele Papo

**Albany Medical Center:** Suzanne Barry, Christopher Woll, Gregory Wu, Erin Carrole, Kathryn Burke, Mustafa Mohammed

**Allina Health (Abbott Northwestern Hospital, United Hospital and Mercy Hospital in Minnesota):** Catherine A. St. Hill, Roman R. Melamed, David M. Tierney, Love A. Patel, Vino S. Raj,Barite U. Dawud, Narayana Mazumder, Abbey Sidebottom, Alena M. Guenther, Benjamin D. Krehbiel, Nova J. Schmitz, Stacy L. Jepsen

**AnMed Health:** Abhijit A Raval, Andrea Franks

**Arkansas Children’s Hospital:** Katherine Irby, Ronald C. Sanders Jr., Glenda Hefley

**Ascension St. Mary’s Hospital:** Jennifer M. Jarvis

**Ascension St**.**Vincent Hospital Indianapolis:** Anmol Kharbanda, Sunil Jhajhria, Zachary Fyffe

**Ascension/St. Thomas Research Institute West Campus:** Bethany Alicie

**Augusta Health:** Andrew S. Moyer, George M. Verghese

**Augusta University Medical Center:** Andrea Sikora Newsome, Christy C. Forehand, Rebecca Bruning, Timothy W. Jones

**Aultman Hospital:** Moldovan Sabov

**Banner University Medical Center-Tucson:** Jarrod M Mosier, Karen Lutrick, Beth Salvagio Campbell, Cathleen Wilson, Patrick Rivers, Jonathan Brinks, Mokenge Ndiva Mongoh, Boris Gilson

**Baylor College of Medicine, Baylor St. Lukes Medical Center:** Christopher M Howard, Cameron McBride, Jocelyn Abraham, Orlando Garner, Katherine Richards, Keegan Collins, Preethi Antony, Sindhu Mathew

**Baylor Scott & White Health:** Valerie C. Danesh, Gueorgui Dubrocq, Amber L. Davis, Marissa J Hammers, ill M. McGahey, Amanda C. Farris, Elisa Priest, Robyn Korsmo, Lorie Fares, Kathy Skiles, Susan M. Shor, Kenya Burns, Corrie A Dowell, Gabriela “Hope” Gonzales, Melody Flores, Lindsay Newman, Debora A Wilk, Jason Ettlinger, Jaccallene Bomar, Himani Darji, Alejandro Arroliga, Alejandro C Arroliga, Corrie A. Dowell, Gabriela Hope Conzales, Melody Flores, Lindsay Newman, Debora A. Wilk, Jason Ettlinger, Himani Darji, Jaccallene Bomar

**Beth Israel Deaconess Medical Center:** Valerie M. Banner-Goodspeed, Somnath Bose, Lauren E. Kelly, Melisa Joseph, Marie McGourty, Krystal Capers, Benjamin Hoenig, Maria C. Karamourtopoulos, Anica C. Law, Elias N. Baedorf Kassis

**Boston University School of Medicine, Boston, MA:** Allan J. Walkey, Sushrut S. Waikar, Michael A. Garcia, Mia Colona, Zoe Kibbelaar, Michael Leong, Daniel Wallman, Kanupriya Soni, Jennifer Maccarone, Joshua Gilman, Ycar Devis, Joseph Chung, Munizay Paracha, David N. Lumelsky, Madeline DiLorenzo, Najla Abdurrahman, Shelsey Johnson

**Brooke Army Medical Center:** Maj Andrew M. Hersh, CPT Stephanie L Wachs, Brittany S. Swigger, CPT Stephanie L Wachs, Capt Lauren A. Sattler, Capt Michael N. Moulton

**Buffalo General Medical Center, Millard Fillmore Suburban Hospital and Oishei Children’s Hospital:** Kimberly Zammit, Patrick, J, McGrath, William, Loeffler,Maya, R, Chilbert

**Cardinal Glennon Children’s Hospital:** Aaron S. Miller, Edwin L. Anderson, Rosemary Nagy, Ravali R. Inja

**Cedars Sinai Medical Center:** Pooja A. Nawathe, Isabel Pedraza, Jennifer Tsing, Karen Carr, Anila Chaudhary, Kathleen Guglielmino

**Chambersburg Hospital:** Raghavendra Tirupathi, Alymer Tang, Arshad Safi, Cindy Green, Jackie Newell

**Children’s Hospital Colorado, University of Colorado Anschutz Medical Campus:** Katja M. Gist, Imran A Sayed, John Brinton, Larisa Strom

**CHRISTUS Spohn Hospital Corpus Christi - Shoreline:** Joshua J. White, Shani B. Italiya, Salim Surani, Lynn Carrasco

**Clements University Hospital at UT Southwestern Medical Center:** Sreekanth Cheruku, Farzin Ahmed, Christopher Deonarine, Ashley Jones, Mohammad-Ali Shaikh, David Preston, Jeanette Chin

**Cox Medical Center Springfield:** Steven K. Daugherty, Sam Atkinson, Kelly Shrimpton

**Detar Family Medicine residency:** Sidney Ontai, Brian Contreras, MD, Uzoma Obinwanko, Nneka Amamasi, Amir Sharafi

**DeTar Hospital:** Salim Surani, Sidney C. Ontai, Brian Contreras, Daniel Handayan, Jeremy George, Janet Le, Aniruddha Gollapalli, Iqra Qureshi

**DeTar/Texas A&M Family Medicine Residency:** Harish Chandna, Sidney C. Ontai, Amirhossein Sharafi, Iqra Qureshi, MD, Hasan Yasin

**George Washington University:** David P. Yamane, Ivy Benjenk, Nivedita Prasanna

**Hassenfeld Children’s Hospital at NYU Langone:** Heda R. Dapul, Sourabh Verma, Alan Salas, Ariel Daube, Michelle Korn, Michelle Ramirez, Logi Rajagopalan, Laura Santos

**Howard University Hospital Washington:** Norma Smalls Mantey

**Jacobs Medical Center UC San Diego Health – La Jolla:** Atul Malhotra, Abdurrahman Husain, Qais Zawaydeh

**Johns Hopkins School of Medicine:** J.H. Steuernagle

**JPS Health Network:** Steven Q. Davis, Valentina Jovic, Valentina Jovic, Max Masuda, Amanda Hayes

**KCPCRU at Norton Children’s Hospital Louisville:** Melissa Thomas, Sarah Morris, Jennifer Nason

**LifeBridge Health/Sinai and Northwest Hospitals:** Jaime Simon Barnes, Namrata Nag

**Lincoln Medical Center:** Manoj K Gupta, Franscene E. Oulds, Akshay Nandavar

**Loyola University Medical Center:** Yuk Ming Liu, Sarah Zavala, Sarah Zavala, Esther Shim

**M Health-Fairview, University of Minnesota:** Ronald A. Reilkoff, Julia A. Heneghan, Sarah Eichen, Lexie Goertzen, Scott Rajala, Ghislaine Feussom, Ben Tang

**Mayo Clinic Arizona:** Rodrigo Cartin-Ceba, Ayan Sen, Amanda Palacios, Giyth M. Mahdi

**Mayo Clinic Rochester:** Rahul Kashyap, Ognjen Gajic, Aysun Tekin, Amos Lal, John C. O’Horo, Neha N. Deo, Mayank Sharma, Shahraz Qamar

**Mayo Clinic, Florida:** Devang Sanghavi, Pramod Guru, Karthik Gnanapandithan, Hollie Saunders, Zachary Fleissner, Juan Garcia, Alejandra Yu Lee Mateus, Siva Naga Yarrarapu

**Mayo Clinic, Mankato:** Syed Anjum Khan, Juan Pablo Domecq, Nitesh Kumar Jain, Thoyaja Koritala

**Mayo Clinic, Eau Claire**: Abigail T. La Nou, Marija Bogojevic

**Medical Center Navicent Health:** Amy B. Christie, Dennis W. Ashley, Rajani Adiga

**Medical College of Wisconsin:** Rahul S Nanchal, Paul A Bergl, Jennifer L Peterson

**Mercy Hospital and Medical Center, Chicago:** Travis Yamanaka, Nicholas A. Barreras, Michael Markos, Anita Fareeduddin, Rohan Mehta

**Mercy Hospital, Saint Louis:** Chakradhar Venkata, Miriam Engemann, Annamarie Mantese

**Nazareth Hospital Trinity Health Mid-Atlantic Philadelphia:** Racheal Park

**New Hanover Regional Medical Center:** Nasar A Siddiqi, Lesly Jurado, Lindsey Tincher, Carolyn Brown

**OSF Saint Francis Medical Center:** Bhagat S. Aulakh, Sandeep Tripathi, Jennifer A. Bandy, Lisa M. Kreps, Dawn R. Bollinger, Jennifer A. Bandy

**OU Medical Center:** Neha Gupta, Brent R Brown, Tracy L Jones, Kassidy Malone, Lauren A Sinko, Amy B Harrell, Shonda C Ayers, Lisa M Settle, Taylor J Sears

**Parkview Health System, Fort Wayne:** Roger Scott Stienecker, Andre G. Melendez, Tressa A. Brunner, Sue M Budzon, Jessica L. Heffernan, Janelle M. Souder, Tracy L. Miller, Andrea G. Maisonneuve

**Ridgecrest Regional Hospital:** Victoria Schauf

**Roper St. Francis Healthcare Charleston:** Sara Utley, Holly Balcer

**Saint Alphonsus Regional Medical Center:** Kerry P. J. Pulver, Jennifer Yehle, Alicia Weeks, Terra Inman

**Saint Luke’s Hospital:** Majdi Hamarshi, Jeannette Ploetz, Nick Bennett, Kyle Klindworth, Moustafa Younis, Adham Mohamed

**Samaritan Health Services:** Brian L. Delmonaco, Anthony Franklin, Mitchell Heath

**Santa Barbara Cottage Hospital:** Diane Barkas

**Sarasota Memorial Hospital:** Antonia L. Vilella, Sara B. Kutner, Kacie Clark, Danielle Moore

**Seattle Children’s Hospital:** Shina Menon, John K McGuire, Deana Rich

**St. Joseph Mercy Ann Arbor, Ann Arbor:** Harry L. Anderson, III, Dixy Rajkumar, Ali Abunayla, Jerrilyn Heiter

**St. Joseph’s Candler Health System:** Howard A. Zaren, Stephanie J. Smith, Grant C. Lewis, Lauren Seames, Cheryl Farlow, Judy Miller, Gloria Broadstreet

**St**.**Agnes Hospital:** Anthony Martinez, Micheal Allison, Aniket Mittal, Rafael Ruiz, Aleta Skaanland, Robert Ross

**St**.**Mary Medical Center, Langhorne:** Umang Patel, Jordesha Hodge, KrunalKumar Patel, Shivani Dalal, Himanshu Kavani, Sam Joseph

**Stamford Health:** Michael A. Bernstein, Ian K. Goff, Matthew Naftilan, Amal Mathew, Deborah Williams, Sue Murdock, RN, Maryanne Ducey, Kerianne Nelson

**Stanford Hospital and Clinics:** Paul K Mohabir, Connor G O’Brien, Komal Dasani

**SUNY Upstate Medical University:** William Marx, Ioana Amzuta, Asad J. Choudhry, Mohammad T. Azam

**Temple University:** Daniel A Salerno

**The Children’s Hospital at OU Medicine:** Neha Gupta, Tracy L Jones, Shonda C Ayers, Amy B Harrell, Dr. Brent R Brown

**The Children’s Hospital of San Antonio, Baylor College of Medicine:** Utpal S. Bhalala, Joshua Kuehne, Melinda Garcia, Morgan Beebe, Heather Herrera

**The Queen’s Medical Center:** Chris Fiack, Stephanie Guo, May Vawer, Beth Blackburn

**Thomas Jefferson University Hospital:** Katherine A. Belden, Michael Baram, Devin M. Weber, Rosalie DePaola, Yuwei Xia, Hudson Carter, Aaron Tolley

**Truman Medical Centers**: Mark Steele, Laurie Kemble

**Tulane University Medical Center and University Medical Center New Orleans:** Joshua L. Denson, A. Scott Gillet, Margo Brown, Rachael Stevens, Andrew Wetherbie, Kevin Tea, Mathew Moore

**UC San Diego Medical Center – Hillcrest:** Abdurrahman Husain, Atul Malhotra, Qais Zawaydeh

**UC San Diego Medical Center -Jacobs:** Atul Malhotra, Abdurrahman Husain, Qais Zawaydeh

**UNC Medical Center:** Benjamin J Sines, Thomas J Bice

**University Medical Center (University Medical Center of Southern Nevada Las Vegas):** Rajany V. Dy, Alfredo Iardino, Jill Sharma, Richard Czieki, Julia Christopher, Ryan Lacey, Marwan Mashina,, Kushal Patel

**University of Alabama at Birmingham:** Erica C. Bjornstad, Nancy M. Tofil, Scott House, Isabella Aldana

**University of Arkansas for Medical Sciences:** Nikhil K. Meena, Jose D. Caceres, Nikhil K Meena, Sarenthia M. Epps, Harmeen Goraya, Kelsey R. Besett, MD, Ryan James, Lana Y. Abusalem, Akash K. Patel, Lana S Hasan

**University of Chicago:** Casey W Stulce, Grace Chong, Ahmeneh Ghavam, Anoop Mayampurath

**University of Cincinnati:** Dina Gomaa B.S., Michael Goodman, Devin Wakefield, Anthony Spuzzillo, John O. Shinn II

**University of Colorado Hospital:** Robert MacLaren

**University of Florida Health Shands Hospital:** Azra Bihorac, Tezcan Ozrazgat Baslanti, George Omalay, Haleh Hashemighouchani, Julie S. Cupka, Matthew M Ruppert

**University of Iowa Carver College of Medicine:** Patrick W. McGonagill, Colette Galet, Janice Hubbard, David Wang, Lauren Allan, Aditya Badheka, Madhuradhar Chegondi

**University of Kansas Medical Center:** Usman Nazir, Garrett Rampon, Jake Riggle, Nathan Dismang

**University of Louisville Hospital:** Ozan Akca, Rainer Lenhardt, Rodrigo S. Cavallazzi, Ann Jerde, Alexa Black, Allison Polidori, Haily Griffey, Justin Winkler, Thomas Brenzel

**University of Miami Miller School of Medicine:** Roger A. Alvarez, Amarilys Alarcon-Calderon, Marie Anne Sosa, Sunita K. Mahabir, Mausam J. Patel

**University of Michigan Health System:** Pauline Park, Andrew Admon, Sinan Hanna, Rishi Chanderraj, Maria Pliakas, Ann Wolski, Jennifer Cirino

**University of Missouri, Columbia:** Dima Dandachi, Hariharan Regunath, Maraya N. Camazine, Grant. E. Geiger, Abdoulie O. Njai, Baraa M. Saad

**University of Utah Health:** Joseph E. Tonna, Nicholas M. Levin, Kayte Suslavich, Rachel Tsolinas, Zachary T. Fica, Chloe R. Skidmore

**University of Vermont Larner College of Medicine:** Renee D. Stapleton, Anne E. Dixon, Olivia Johnson, Sara S. Ardren, Stephanie Burns, Anna Raymond, Erika Gonyaw, Kevin Hodgdon, Chloe Housenger, Benjamin Lin, Karen McQuesten, Heidi Pecott-Grimm, Julie Sweet, Sebastian Ventrone

**Valleywise Health (formerly Maricopa Medical Center):** Murtaza Akhter, Rania Abdul Rahman, Mary Mulrow

**Vanderbilt University Medical Center:** Erin M. Wilfong, Kelsi Vela

**Wake Forest University School of Medicine; Wake Forest Baptist Health Network:** Ashish K. Khanna, Lynne Harris, Bruce Cusson, Jacob Fowler, David Vaneenenaam, Glen McKinney, Imoh Udoh, Kathleen Johnson

**Washington University School of Medicine and Barnes-Jewish Hospital:** Patrick G. Lyons, Andrew P Michelson, Sara S. Haluf, Lauren M. Lynch, Nguyet M. Nguyen, Aaron Steinberg

**West Virginia University Morgantown:** Ankit Sakhuja

**William S. Middleton Memorial VA Hospital Madison:** Nicholas A Braus

**Yale New Haven Health New Haven:** Kevin N Sheth, Abdalla A Ammar

